# A time-varying risk assessment framework for *P. vivax* malaria transmission in temperate settings: A case study of the Republic of Korea

**DOI:** 10.64898/2026.07.20.26358215

**Authors:** Woo-Sik Son, Ah-Young Lim, Dong-Uk Hwang, Kyeongah Nah

## Abstract

Understanding the time-varying transmission potential of *Plasmodium vivax* is essential for guiding elimination efforts, particularly in low-transmission, temperate regions where the seasonal vector activity and the complex biology of the parasite present unique challenges. Here we develop an integrated modelling approach to estimate the time-varying case reproduction number of *P. vivax* malaria in the Republic of Korea, using weekly data on temperature, malaria activity index, and symptom onset from the primary endemic regions between 2013 and 2025. The model integrates temperature-dependent and seasonally constrained serial intervals to reconstruct the transmission timeline and infer likely transmission links between cases without requiring phylogenetic or contact-tracing data. Our findings reveal that although the greatest number of secondary cases is generated during the summer transmission months (June to August), the average case reproduction number is highest during the pre-transmission season (October to April), indicating that cases arising in the pre-transmission season has a higher potential to generate further infections compared to cases in other seasons. This underscores the need for enhanced case detection, diagnosis, and treatment during the pre-transmission period. To make these patterns explorable, we provide a web-based interactive tool that links each case to the infections it seeds, revealing transmission that crosses years. This modelling framework relies only on routinely collected surveillance and environmental data, offering a transferable tool for resource-constrained or pre-elimination settings where genetic or contact-tracing data are unavailable.

## Introduction

Efforts to eliminate malaria are progressing globally, yet *Plasmodium vivax (P. vivax)* remains a persistent challenge, particularly in countries approaching elimination [1]. Unlike *Plasmodium falciparum (P. falciparum)*, which predominates in high-burden tropical settings, *P. vivax* thrives in temperate zones, where transmission is confined to the warmer months when *Anopheles* vectors are active [2]. The biology of the parasite further complicates control: dormant liver-stage hypnozoites can reactivate months or years after the initial infection, blood-stage parasitaemia is often too low for routine microscopy to detect, and the intrinsic incubation period ranges from weeks to over a year [3–5]. These features allow *P. vivax* to sustain low-level transmission that evades routine surveillance, even in settings with strong health infrastructure [4, 5]. As countries progress towards elimination, identifying which periods contribute most to onward transmission becomes essential, yet requires analytical approaches capable of capturing the pronounced seasonality and complex temporal dynamics that characterise *P. vivax* malaria in temperate regions [6, 7].

The case reproduction number, *R*_*t*_, defined as the expected number of secondary cases generated by a single case with symptom onset at time *t* [8], is widely used to assess time-varying transmission intensity. For directly transmitted infections, estimation of *R*_*t*_ relies on a serial interval distribution that depends only on the elapsed time between successive symptom onsets [8]; for vector-borne diseases, however, the serial interval spans a host–vector–host cycle shaped by temperature and mosquito activity [9, 10]. Temperature-dependent generation intervals have been modelled for *P. falciparum* [9] and dengue [10], and renewal equation frameworks have been applied to *P. vivax* malaria in the Republic of Korea to account for its bimodal incubation period and the restriction of transmission to the mosquito-active season [11]. Yet for *P. vivax* transmission in temperate settings, a further constraint arises: the mosquito-mediated steps of the transmission cycle can occur only when vectors are active, while the parasite can persist in the human host through winter. The serial interval therefore depends not only on the elapsed time between symptom onsets but on when in the seasonal cycle the infector’s symptoms begin: a case arising when mosquitoes are inactive faces a serial interval of many months, whereas a case arising at the height of the mosquito season can complete a transmission cycle within weeks. No existing framework for *P. vivax* malaria constructs serial intervals that vary explicitly with the infector’s symptom onset time while jointly integrating empirical temperature records, seasonal mosquito activity, and the bimodal incubation period. Developing and validating such an approach requires a setting with longitudinal case surveillance and concurrent epidemiological and environmental monitoring.

The Republic of Korea provides a well-characterized setting in which to develop such a framework. After achieving elimination in the late 20th century, the country experienced a reintroduction of *P. vivax* malaria along the Demilitarized Zone in 1993, and transmission has since remained largely confined to the northern border regions of Seoul, Incheon, Gyeonggi, and Gangwon [12]. Despite sustained control efforts, the national goal of elimination by 2030 remains unmet, with progress constrained by the biological complexity of *P. vivax* and the persistence of low-level transmission [12, 13]. The national surveillance system provides weekly case notifications, complemented by daily temperature records from 23 meteorological stations and a weekly mosquito vector index across the endemic regions. These concurrent data streams offer the temporal resolution needed to construct environmentally informed serial intervals and to examine how transmission intensity varies within and between years.

Here we develop a framework in which the serial interval distribution varies explicitly with the calendar time of the infector’s symptom onset. The model integrates temperature-dependent parasite development within the mosquito, seasonal variation in mosquito activity, and the bimodal intrinsic incubation period of *P. vivax* into a unified construction of the serial interval. Applying this framework to weekly case, temperature, and vector surveillance data from 2013 to 2025, we estimate the time-varying case reproduction number and decompose transmission into within-season and cross-season components to identify the periods that contribute most to onward transmission. To support interpretation of these seasonal patterns, we provide a publicly accessible interactive visualisation that resolves, for cases arising in any chosen period, when the secondary infections they generate subsequently develop symptoms. By relying on routinely collected epidemiological and environmental data, this framework provides a basis for assessing seasonal transmission intensity in temperate *P. vivax* transmission settings where case counts alone offer limited insight into the timing and structure of onward transmission.

## Methods

### Data

To construct serial interval distributions and estimate the time-varying case reproduction number, we assembled three routinely collected datasets covering the four upper-level administrative areas that account for most malaria transmission in the Republic of Korea: Seoul, Incheon, Gyeonggi, and Gangwon.

We obtained weekly counts of *P. vivax* malaria cases from the Infectious Disease Portal of the Korea Disease Control and Prevention Agency (KDCA) [14]. For each week, we summed the locally acquired cases reported across the four study regions and excluded imported cases, yielding a weekly time series of locally acquired infections from 2013 to 2025.

We obtained daily mean temperatures from the open data portal of the Korea Meteorological Administration [15]. For each day, we averaged the mean temperatures recorded at the 23 Automated Synoptic Observing System (ASOS) stations in the four study regions to produce a single daily temperature series from 2013 to 2025, used to drive the temperature-dependent extrinsic incubation period (EIP).

Mosquito activity was quantified by the malaria vector index published in the KDCA’s annual guidelines for the surveillance and management of malaria vector mosquitoes [16–28]. The malaria vector index for a given week is defined as the number of female *Anopheles* mosquitoes collected per trap-night using black-light traps operated from 18:00 to 06:00. KDCA conducts monitoring annually during weeks 14 to 44 as part of the national vector surveillance program; the first and last weeks of this window typically record zero or near-zero values, capturing the onset and cessation of vector activity each year. Because the index was recorded at integer precision before 2022, small positive values during low-activity weeks were entered as zero; we replaced these zeros within the monitoring period with a small positive value estimated from regional surveillance records (S1 Text).

All three datasets, together with the complete analysis code, are publicly available at the study repository (see Data and code in Acknowledgments)

### Time-varying case reproduction number

To identify the periods that contribute most to onward transmission of *P. vivax malaria*—and therefore when case detection and treatment should be prioritized to prevent secondary infections—we estimated the time-varying case reproduction number. For an individual infected at time *t*, the case reproduction number is the expected number of secondary cases that this individual subsequently generates [8, 29]; in the context of malaria, it counts the secondary human infections seeded by a case whose symptoms begin at time *t*, via the mosquito vector. Because the timing of infection is not directly observed, we use the symptom onset time series in its place throughout.

The case reproduction number at time *t, R*_*t*_, is the mean of *R*_*j*_, which is the expected number of secondary cases generated by a case *j* whose symptom began at time *t*. It is obtained as the sum, over every case *i* that developed symptoms after *j*, of the likelihood *p*_*ij*_ that *i* was infected by *j* [8]:

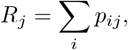

Computing this likelihood requires the probability distribution of the serial interval. We write *w*_*x,t*_ for the probability that the serial interval—the time from an infector’s symptom onset to the corresponding infectee’s symptom onset—is *x* when the infector’s symptoms begin at time *t*. The case reproduction number was first introduced for severe acute respiratory syndrome (SARS) [8], where the serial interval distribution *w*_*x*_ depends only on the difference *x* between the infector’s and infectee’s symptom onset times, regardless of when the infector developed symptoms. For *P. vivax* malaria in a temperate setting, however, the serial interval is strongly shaped by the calendar timing of transmission: mosquito activity is highly seasonal, and the development of the parasite inside the mosquito is temperature-dependent. We therefore adapt the case reproduction number method specifically for *P. vivax* transmission by allowing the serial interval distribution *w*_*x,t*_ to depend on the infector’s symptom onset time *t* itself, rather than on the onset-time difference alone.

The infection likelihood then becomes

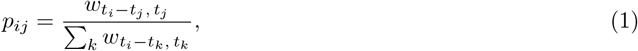

where *t*_*i*_, *t*_*j*_, and *t*_*k*_ represent the symptom onset times of cases *i, j*, and *k*, respectively.

Using the estimated *R*_*t*_ values, we then summarized the absolute volume of onward transmission by season. Specifically, we computed the expected number of secondary cases for each week as the product of *R*_*t*_ and the number of cases with symptom onset in that week. Weekly values were then averaged within each calendar month across all years to obtain the expected number of secondary cases per month.

### Serial interval model

The serial interval *x* spans a complete human–mosquito–human transmission cycle and is composed of four successive intervals (Fig 1), following Codeço et al. [10]: the interval from the infector’s symptom onset to the infection of a mosquito (*T*_*HM*_), the EIP during which the parasite develops inside the mosquito (*I*_*M*_), the interval from the mosquito becoming infectious to its infecting a new host (*T*_*MH*_), and the intrinsic incubation period in the newly infected human (*I*_*H*_), so that *x* = *T*_*HM*_ + *I*_*M*_ + *T*_*MH*_ + *I*_*H*_.

**Fig 1.**
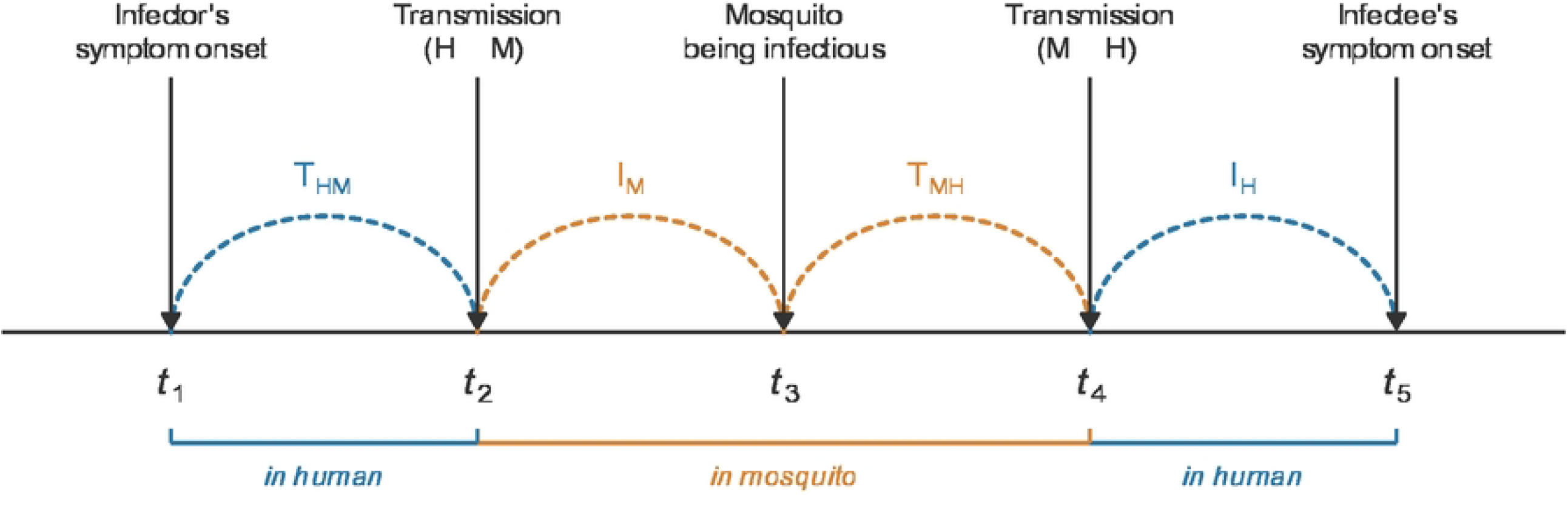
Four components of the serial interval for *P. vivax* malaria. The serial interval *x* spans a complete human–mosquito–human transmission cycle and is the sum of four successive intervals: *T*_*HM*_, from the infector’s symptom onset (*t*_1_) to infection of a mosquito (*t*_2_); *I*_*M*_, the EIP in the mosquito (*t*_2_ to *t*_3_); *T*_*MH*_, from the mosquito becoming infectious to its infecting a new host (*t*_3_ to *t*_4_); and *I*_*H*_, the intrinsic incubation period in the newly infected human, ending at symptom onset (*t*_5_); so that *x* = *T*_*HM*_ + *I*_*M*_ + *T*_*MH*_ + *I*_*H*_. Colored segments denote phases in the human host (blue) and in the mosquito vector (orange).

Let *h*_*HM*_ (*s, t*) be the rate at which a person whose symptoms began *s* time units earlier transmits parasites to a mosquito at time *t*. Because this transmission requires a blood-feeding mosquito, the rate scales with the level of mosquito activity *M* (*t*):

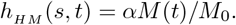

Here *M* (*t*) is quantified by the weekly malaria vector index described in the Data section, and *M*_0_ is a reference peak value set to 12, the median of the annual maximum vector index values across 2013–2025. This median corresponds to the value observed in week 28 of 2017 — so that *M* (*t*)/*M*_0_ = 1 at this reference week. Human-to-mosquito transmission occurs during the blood-stage infection through the ingestion of gametocytes, whose densities decline after one to two months of patency [30]. The probability that an individual whose symptoms began *s* time units earlier has not yet transmitted to a mosquito by time *t* is

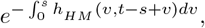

and the probability distribution of *T*_*HM*_, the interval between the infector’s symptom onset (*t*_1_) and transmission to a mosquito (*t*_2_), is

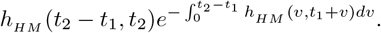

We set *α* to lie between 0.2 and 0.4. At the representative peak week, where *M* (*t*)/*M*_0_ = 1 so that *h*_*HM*_ = *α*, the probabilities that an infected individual has transmitted to a mosquito within 7 and within 14 days of symptom onset, namely 1 − *e*^−7*α*^ and 1 − *e*^−14*α*^, are summarized in Table 1 for *α* = 0.2, 0.3, and 0.4.

**Table 1.** Probability of human-to-mosquito transmission within 7 and 14 days of symptom onset.

| $\alpha$ | Within 7 days | Within 14 days |
| --- | --- | --- |
| 0.2 | 75.3% | 93.9% |
| 0.3 | 87.8% | 98.6% |
| 0.4 | 93.9% | 99.7% |

Starting from *t*_2_, the time of transmission from an infected human to a mosquito, the mosquito undergoes the EIP, which is the time required for the parasites to develop and make the mosquito transmissible. The EIP (*I*_*M*_) of *P. vivax* has been modelled as a function of mean environmental temperature [31, 32]. When the temperature is held constant at c (°C), the mean EIP is estimated to be 105/(*c* − 14.5) days. Using this estimate, we model *h*_*M*_ (*s, t*), the instantaneous rate at which a mosquito becomes infectious at time *t*, which is *s* time units after the mosquito’s infection, as a function of the temperature *T* (*t*) at that time:

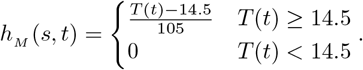

At the representative high-activity week used to normalize the vector index (week 28 of 2017, *M* (*t*)/*M*_0_ = 1), for selected values of *α*.

When the temperature is constant as *T* (*t*) = *c* and *c* > 14.5, the mean EIP is equal to

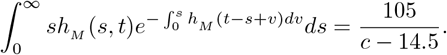

The probability distribution of the EIP (*I*_*M*_) between time *t*_2_ and *t*_3_ is

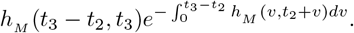

*Anopheles* mosquitoes can overwinter as adults in diapause [33], but the development of *P. vivax* within the mosquito requires temperatures above approximately 16 °C, below which the parasite does not survive [34]; the parasite therefore cannot survive inside an overwintering vector. We therefore require the mosquito’s infection (*t*_2_) and the completion of its EIP (*t*_3_) to fall within the same mosquito-active season. The infected human, by contrast, can carry the parasite through the winter, so the interval from symptom onset (*t*_1_) to the infecting bite (*t*_2_) may span an inactive season; a complete transmission cycle can therefore last longer than a year.

After becoming infectious, the mosquito can transmit the parasite when it next feeds. Let *h*_*MH*_ (*s, t*) be the rate at which this mosquito infects a person at time *t*, a time *s* after it became infectious. This is the same mosquito that fed on the infector at *t*_2_ (Fig 1); for it to bite again at *t*_4_, mosquitoes must still be active, so that *t*_4_ falls within the mosquito-active season (weeks 14–44). Accordingly, *h*_*MH*_ does not depend on the malaria vector index *M* (*t*) but only on whether mosquitoes are active: it vanishes outside the mosquito-active season, when *M* (*t*) = 0, and equals a constant *β* during the season:

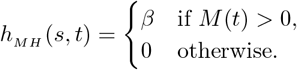

The probability distribution of *T*_*MH*_, the interval from the mosquito becoming infectious (*t*_3_) to its infecting a person (*t*_4_), is

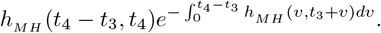

We set *β* to lie between 0.2 and 0.4, jointly with *α*. Over this range, the resulting distribution of *I*_*M*_ + *T*_*MH*_ = *t*_4_ − *t*_2_, the total time the parasite spends within the mosquito, has its peak at 12–13 days (S1 Fig), consistent with the reported lifespan distribution of female *Anopheles sinensis*, the principal *P. vivax* vector in Korea [35, 36]. The primary analyses presented in the main text use *α* = *β* = 0.3.

Using the parameter estimates from [37], the distribution of the intrinsic incubation period is approximated by a linear combination of a Gamma distribution and a log-normal distribution to incubation-period data inferred from the travel history of *P. vivax*-infected cases in the Republic of Korea.

Using this result, we assume that the intrinsic incubation period (*I*_*H*_) between *t*_4_ and *t*_5_ follows the bimodal probability density function

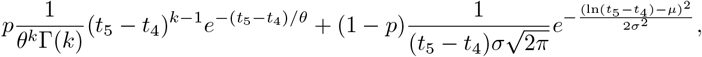

where *p* = 0.7423, *θ* = 1.1141, *k* = 22.8197, *µ* = 5.7851, and *σ* = 0.1410, and Γ denotes the gamma function.

### Constructing the serial interval distribution

The serial interval distribution *w*_*x,t*_ has no closed form, since it arises from the chain of temperature- and season-dependent steps described above; we therefore constructed it numerically. Because the serial interval can extend to as long as two years (Fig 2), left- and right-truncation would bias *R*_*t*_ near the ends of the study period; we therefore restricted estimation of *R*_*t*_ to 2015–2023. For each candidate infector symptom onset time *t*_1_—every week from 2013 to 2025—we drew 1,500,000 sampled sets of (*t*_2_, *t*_3_, *t*_4_, *t*_5_) from the component distributions and recorded the serial interval *x* = *t*_5_ − *t*_1_, building up 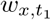 for that onset week.

**Fig 2.**
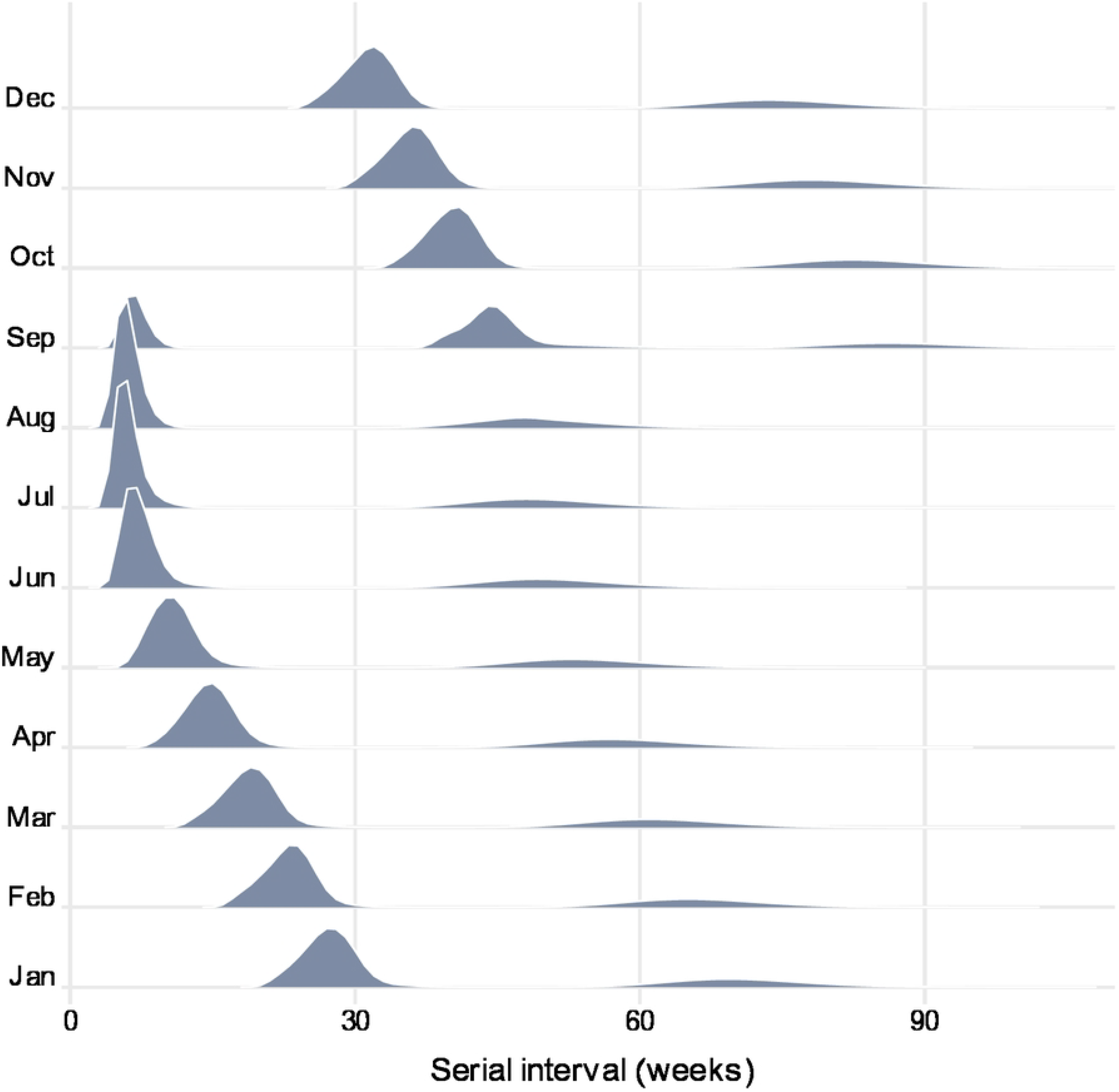
Serial interval distributions by the infector’s month of symptom onset. Each row shows the probability density of the serial interval for infectors whose symptoms began in the indicated month, constructed by Monte Carlo simulation (1,500,000 samples per onset week) of the four-component transmission model (Fig 1).

Two seasonal constraints were enforced by rejection sampling. Because the vector is active only during weeks 14–44 of the year, any set in which *t*_2_ or *t*_4_ (Fig 1) fell outside this window was discarded and resampled. Likewise, because the parasite cannot survive inside an overwintering vector, any set in which the mosquito’s infection (*t*_2_) and the completion of its EIP (*t*_3_) fell in different years was discarded and resampled.

The resulting distribution varies markedly with the infector’s symptom onset time *t*_1_, reflecting mosquito overwintering, the seasonal variation in the vector index *M* (*t*), and the temperature dependence of parasite development inside the mosquito. Fig 2 summarizes this seasonal dependence: for an infector with symptom onset in January, the serial interval shows a first peak at 27 weeks and a second peak at 69 weeks. As the onset month advances, the first peak shortens to a minimum of 6 weeks for an August onset and then rises slightly to 7 weeks in September, while the second peak reaches its minimum of 44 weeks; by October, mosquito overwintering lengthens the intervals again, shifting the first peak to 40 weeks and the second peak to 82 weeks.

Because the serial interval distribution depends so strongly on *t*_1_, the number of secondary cases that a single case is expected to generate likewise varies with its season of symptom onset—the seasonal dependence that the time-varying case reproduction number is designed to capture, and that we quantify in the Results that follow.

## Results

Weekly *R*_*t*_ was estimated for 2015 to 2023 (see Methods). Annual case totals fluctuated between 256 and 581 from 2013 through 2021, then rose to 638 in 2023 and 619 in 2024 (Fig 3A). Weekly *R*_*t*_ followed a seasonal cycle throughout most of the estimation period, rising above and falling below one within each year (Fig 3B). This pattern was interrupted from late 2021: *R*_*t*_ increased sharply and remained continuously above one throughout 2022, before returning to a seasonal cycle in 2023.

**Fig 3.**
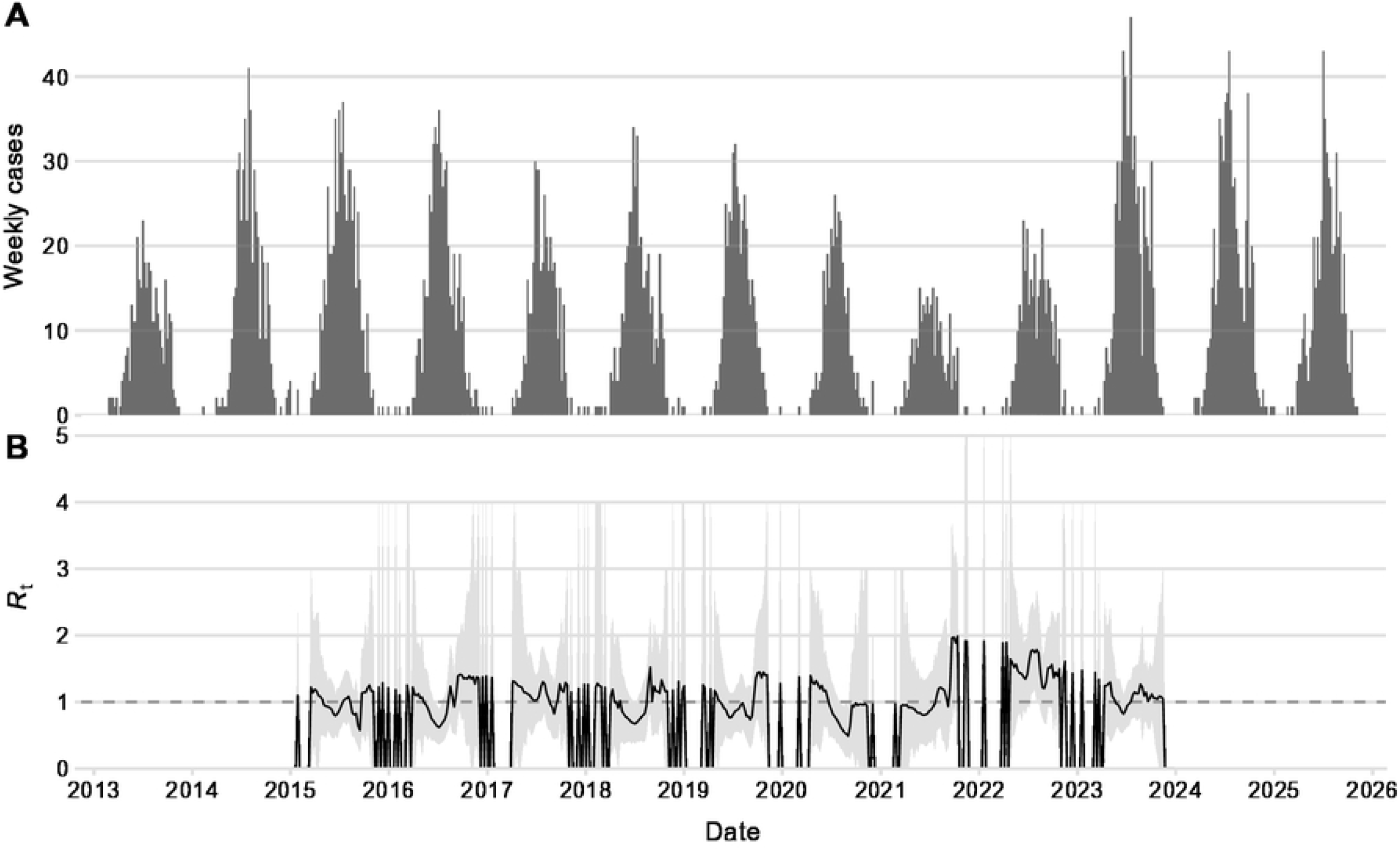
Weekly *P. vivax* malaria cases and time-varying case reproduction number. (A) Weekly counts of locally acquired *P. vivax* malaria cases summed across Seoul, Incheon, Gyeonggi, and Gangwon, 2013–2025. (B) Estimated weekly case reproduction number *R*_*t*_ (black line) with 95% uncertainty intervals (grey shading). *R*_*t*_ was estimated for 2015–2023 to avoid truncation bias (see Methods); weeks with no reported cases yield *R*_*t*_ = 0. The dashed horizontal line marks *R*_*t*_ = 1. All estimates use *α* = *β* = 0.3.

The monthly mean *R*_*t*_, averaged across all years, was highest in January (1.36) and lowest in June (0.93, below one), declining through spring and rising again over the second half of the year (Fig 4, left axis). The expected number of secondary cases per month followed the opposite pattern, peaking in July (23.4) and reaching its lowest values during the mosquito-inactive months (Fig 4, right axis). Although per-case *R*_*t*_ was highest during the mosquito-inactive season, the absolute number of secondary infections was greatest from June to August, when incidence was highest.

**Fig 4.**
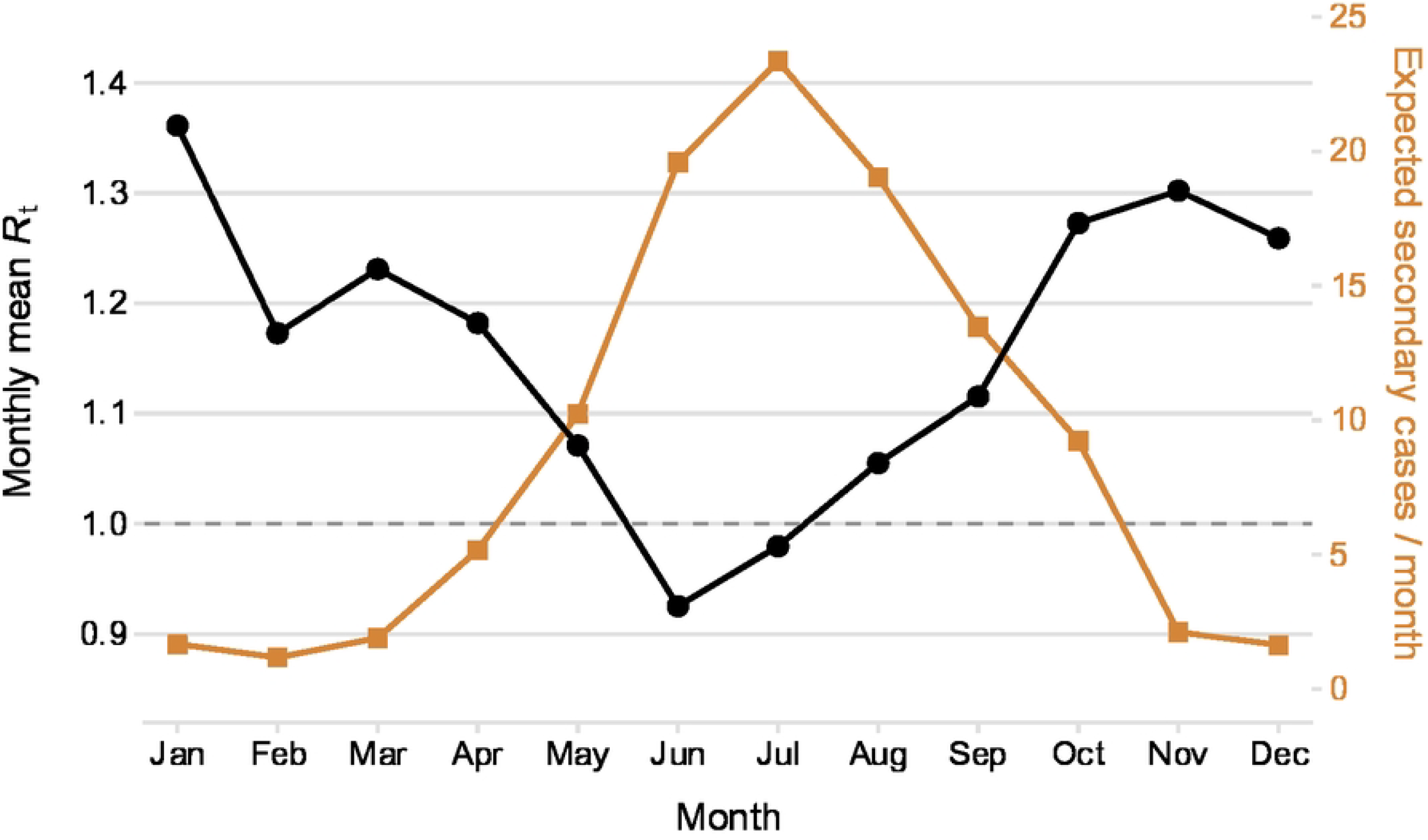
Seasonal profiles of per-case transmission potential and absolute secondary case burden. For each calendar month, averaged across 2015–2023: the mean *R*_*t*_ (black line, left axis) and the expected number of secondary cases per month (orange lines, right axis). The expected number of secondary cases per month is computed as the weekly product of *R*_*t*_ and the number of cases with symptom onset in that week, averaged within each calendar month (see Methods). The dashed horizontal line marks *R*_*t*_ = 1. All estimates use *α* = *β* = 0.3.

The annual mean *R*_*t*_ provides a year-level summary of these dynamics (Fig 5). The annual mean remained between 0.96 and 1.15 from 2015 through 2021, exceeding one in most years and falling below one only in 2020, a year in which COVID-19-related disruptions may have affected healthcare-seeking behavior and case detection. The annual mean rose to its highest value of 1.54 in 2022, coinciding with the sustained elevation in weekly *R*_*t*_ (Fig 3B), before declining to 1.10 in 2023. Estimates were consistent when *α* and *β* were varied independently across their full range (S2 Fig, S3 Fig).

**Fig 5.**
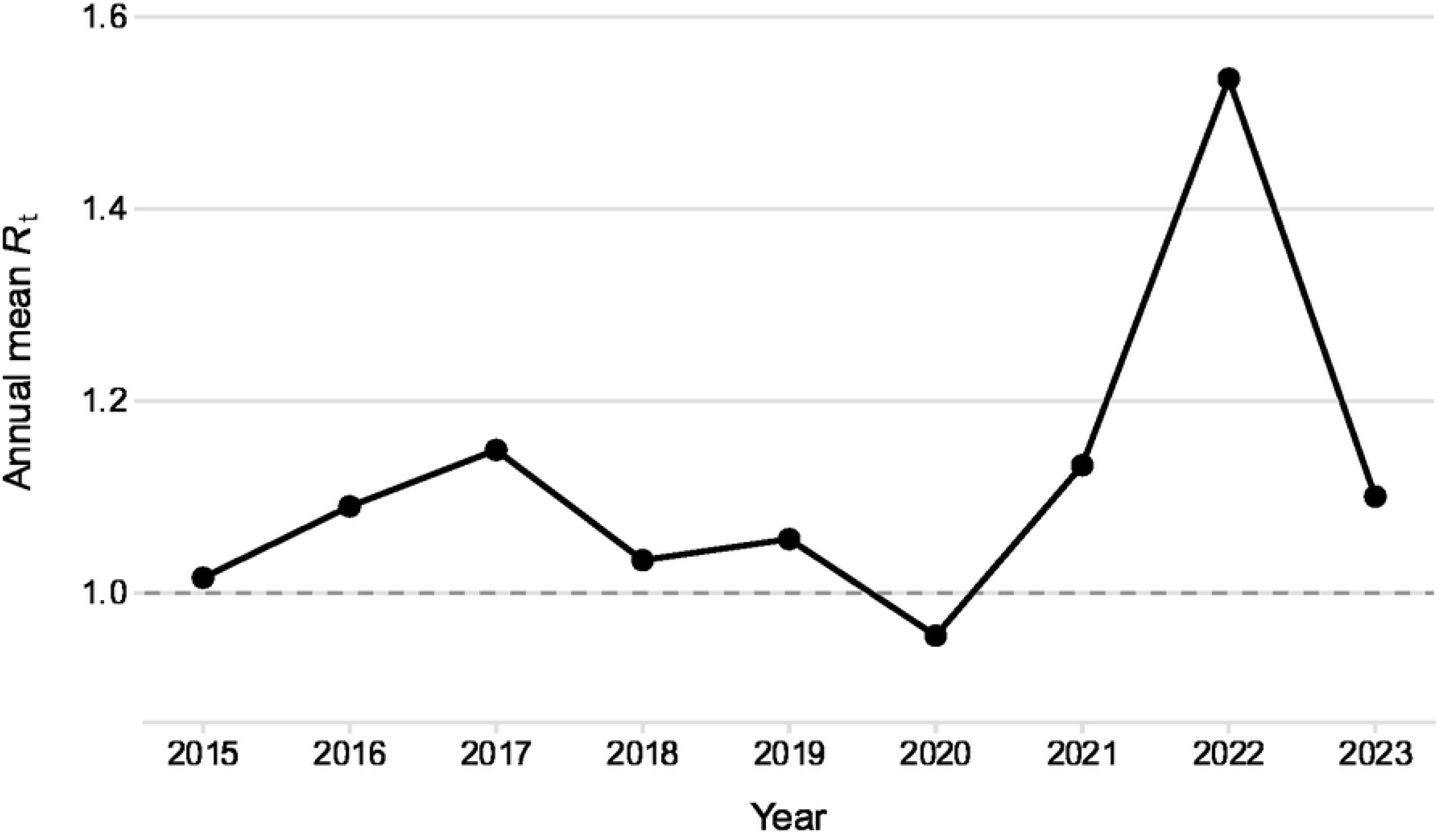
Annual mean case reproduction number, 2015–2023. Each point shows the mean of weekly *R*_*t*_ within a calendar year. The dashed horizontal line marks *R*_*t*_ = 1. All estimates use *α* = *β* = 0.3.

Stratifying inferred transmission links into same-year and cross-year components revealed the temporal structure underlying this seasonal contrast (Fig 6). Among infectees with symptom onset before June, the majority were linked to an infector from the previous year (Fig 6A), indicating that early-season cases were predominantly seeded by the preceding transmission season. Among infectors, the cross-year fraction rose progressively from late summer and reached 100% for those with onset from late September onward (Fig 6B); all secondary cases attributed to these late-season infectors developed symptoms the following year. As observed, mid-summer infectors predominantly generate same-year secondary cases; however, despite their large volume, their per-case transmission potential remains low because subsequent environmental conditions and vector activity progressively decline, truncating their ongoing transmission pathways. In contrast, autumn and winter-onset infectors exclusively generate cross-year secondary cases. This small cohort of off-season infectors monopolizes the transmission into the subsequent year, resulting in the elevated *R*_*t*_.

**Fig 6.**
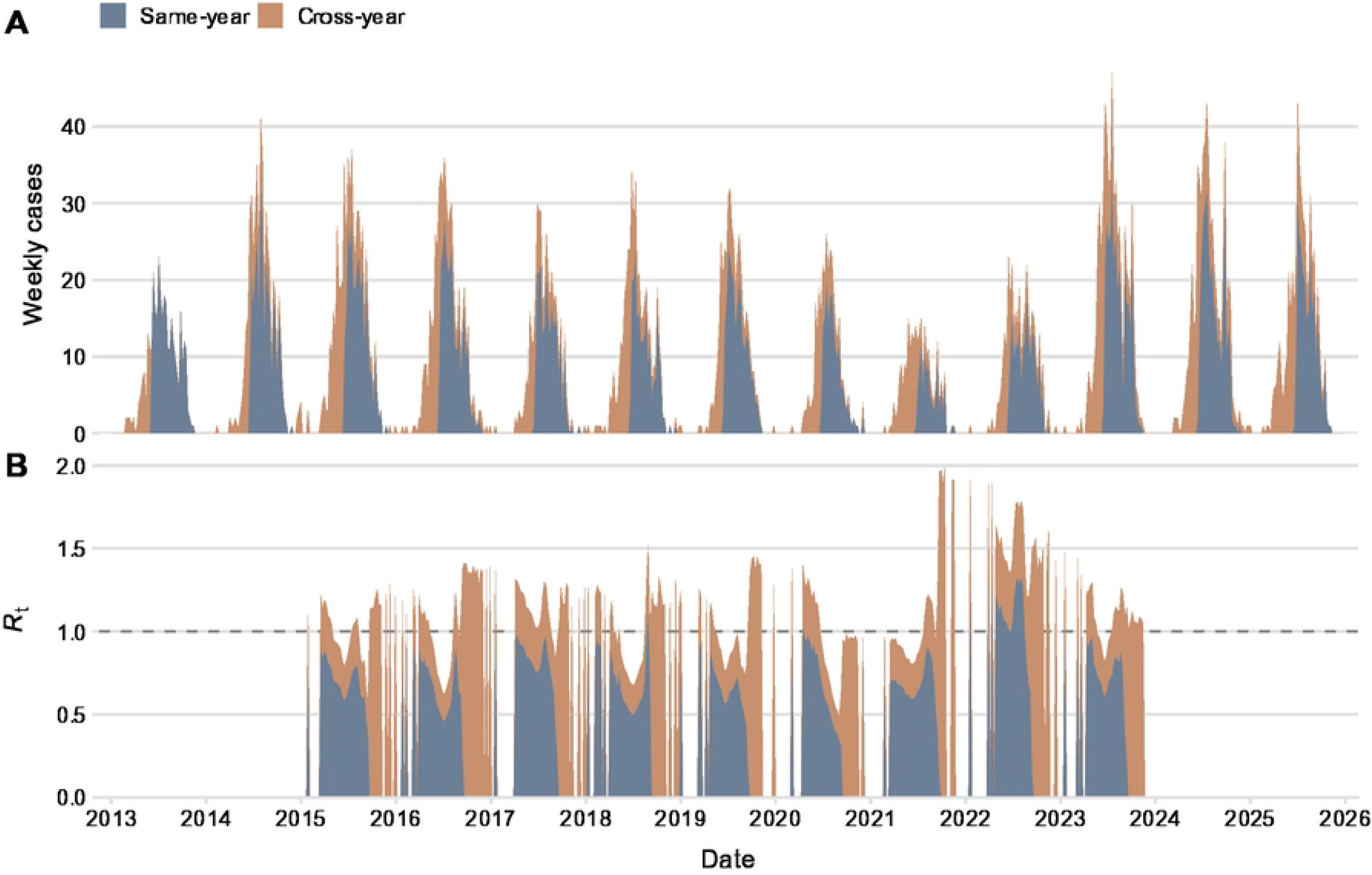
Same-year and cross-year composition of transmission links. Each inferred transmission link is classified by whether infector and infectee developed symptoms in the same calendar year (same-year, blue) or in different years (cross-year, orange). (A) For infectees grouped by week of symptom onset: proportion whose inferred infector developed symptoms in the same year (blue) versus the previous year (orange). (B) For infectors grouped by week of symptom onset: proportion of their inferred infectees who developed symptoms in the same year (blue) versus the following year (orange). Panel A thus shows where each season’s cases came from; panel B shows where each season’s cases transmit to. All estimates use *α* = *β* = 0.3.

Building on this same-year/cross-year decomposition, we developed an interactive visualization that maps the expected symptom onset times of secondary cases arising from individuals infected during a given period, as demonstrated in Fig 7. Cases selected from a summer peak (colored points, Fig 7A) were linked to secondary cases whose symptom onsets separated into two clusters: one within the same season and a second the following year, consistent with the bimodal serial interval (Fig 2; Fig 7B). An interactive version of this visualization, allowing users to select any onset period, is available at https://epirt.notion.site/malaria-rt.

**Fig 7.**
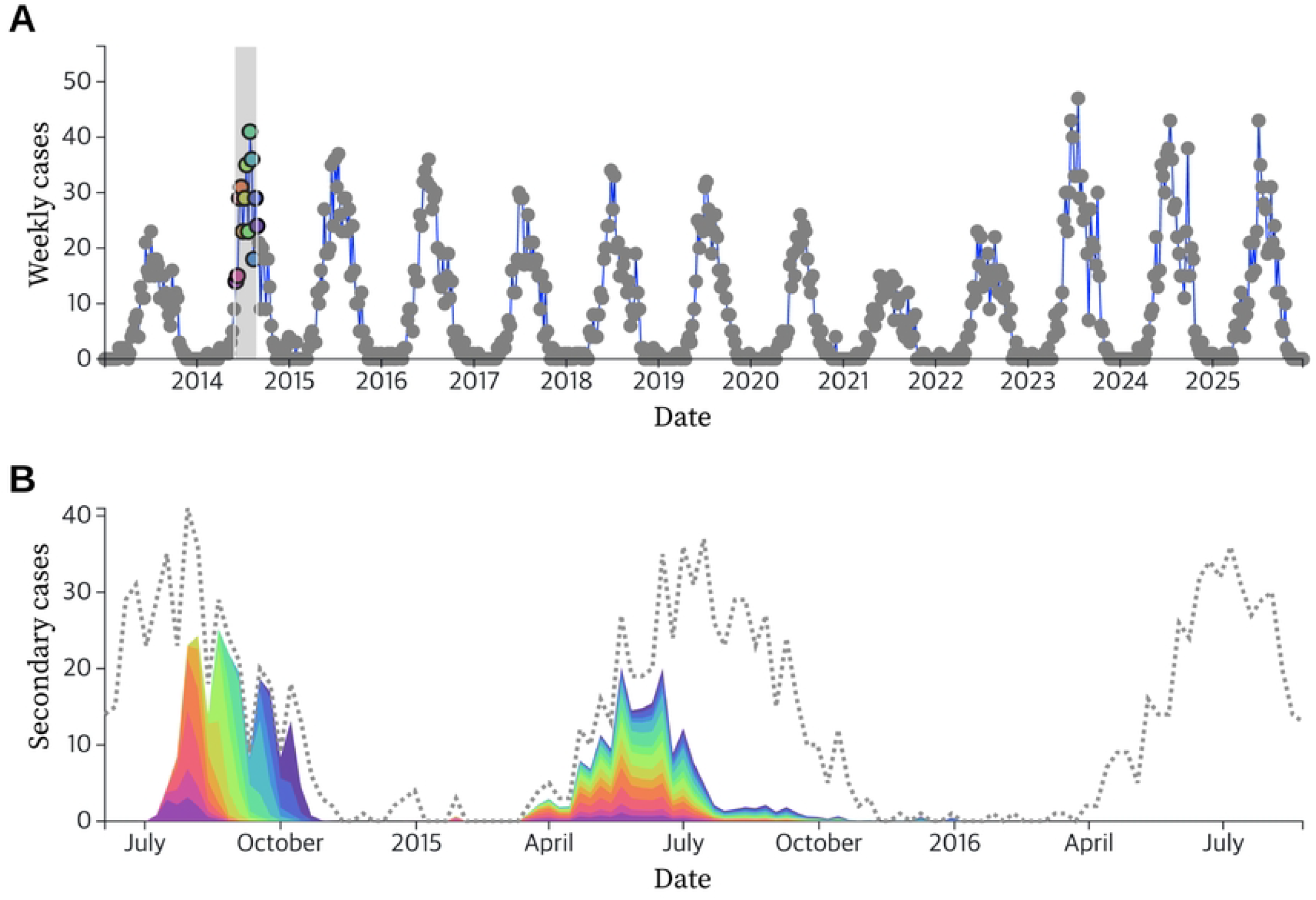
Onset-time distribution of secondary cases attributed to selected infectors. (A) Weekly case counts with a user-selected subset highlighted as colored points. (B) For the selected cases in (A), the onset-time distribution of the secondary cases attributed to them, shown as a stacked series with colors matching the selected points. In the example shown, cases selected from a summer peak are linked to secondary cases whose onsets fall into two clusters: one within the same season and a second the following year. An interactive version allowing selection of any onset period is available at https://epirt.notion.site/malaria-rt.

## Discussion

Here we develop a framework in which the serial interval distribution for *P. vivax* malaria varies with the calendar time at which an infector develops symptoms, rather than depending only on the elapsed time to the infectee. Applying this framework to 13 years of surveillance data from the Republic of Korea, we find that per-case transmission potential is highest not during the summer months when most secondary infections occur, but during the preceding pre-transmission period when mosquitoes are inactive and the transmission cycle cannot resume for months. This contrast indicates that the timing of case detection matters, because infections detected before the mosquito season may carry disproportionate onward transmission potential once transmission becomes possible again.

This seasonal pattern in per-case *R*_*t*_ extends earlier work by Endo and Nishiura [11], who estimated yearly reproduction numbers for *P. vivax* malaria in the Republic of Korea from 1993 to 2012 but averaged over within-year variation. At the weekly scale, we find that *R*_*t*_ exceeds one predominantly during the months when mosquitoes are inactive, a pattern driven by the small number of baseline cases at that time whose subsequent transmission is delayed until the next available transmission window. This is analogous to findings in dengue, where Codeço et al. [10] showed that temperature-dependent generation intervals shift *R*_*t*_ estimates relative to fixed-interval approaches, though in tropical settings the effect is continuous rather than gated by a discrete inactive season. The timing of mosquito activity thus shapes not only aggregate *R*_*t*_ but also the distribution of transmission pathways within and across seasons.

The mechanism underlying this seasonal *R*_*t*_ pattern is supported by the predicted shift from long- to short-incubation cases across the year (S4 Fig), which is independently consistent with the travel-history-based distribution reconstructed by Kim et al. [38]. The cross-year decomposition further reveals that nearly all cases with onset before June trace back to the preceding transmission season, while cases arising from late September onward seed infections exclusively in the following year (Fig 6). To our knowledge, this year-crossing structure has not previously been characterized for *P. vivax* malaria in the Republic of Korea. It clarifies why the pre-transmission months carry the elevated per-case *R*_*t*_, as these cases have access to a full upcoming mosquito season in which to generate secondary infections.

These findings identify the pre-transmission months as a priority window for strengthening case detection in support of the Republic of Korea’s goal of malaria elimination by 2030 [13]. Strengthening surveillance and accelerating diagnosis during the pre-transmission period could yield a greater reduction in onward transmission per case managed than equivalent efforts during the peak season, when per-case *R*_*t*_ is lower. The documented average delay of 4.75 days between symptom onset and diagnosis in the Republic of Korea [39] underscores the potential gains, as the consequences of delayed detection for onward transmission are amplified when per-case *R*_*t*_ is at its seasonal maximum. Such seasonal prioritization of case detection would complement existing summer-focused vector control, together covering both the period of highest absolute transmission and the period of highest per-case transmission potential. Beyond seasonal targeting, the model predicts serial intervals extending beyond two years because infections acquired late in one transmission season may not lead to symptomatic onward cases until after one or more subsequent mosquito seasons (Fig 2), underscoring the importance of sustained case surveillance following the apparent interruption of local transmission.

Our framework does not account for transmission arising from late hypnozoite relapses. Reported relapse rates following treatment in the Republic of Korea are low [40], although even infrequent relapses during the pre-transmission season could carry high per-case transmission potential under this framework. Other factors that may influence the intrinsic incubation period, including parasite inoculum size and treatment regimen, were not incorporated and would require patient-level data to explore. The mosquito-to-human transmission component assumes a constant biting rate during the active season and does not explicitly model mosquito lifespan or age-dependent mortality, simplifications that may underestimate temporal variation in vectorial capacity. Nevertheless, the biting parameters were calibrated so that the time the parasite spends within the vector is consistent with the reported lifespan of *Anopheles* sinensis (S1 Fig). Similarly, the regional temperature series used to drive EIP is a spatial average across 23 stations spanning lowland and highland areas; finer-resolution temperature data could improve local estimates of parasite development rates. These simplifications were pragmatic given the available data, and the robustness of *R*_*t*_ to variation in *α* and *β* (S2 Fig, S3 Fig) suggests that the seasonal patterns reported here are governed primarily by the timing of mosquito activity and temperature rather than by the assumed rate values.

This study provides the first estimates of a weekly case reproduction number for *P. vivax* malaria that accounts for the calendar-time dependence of the serial interval in a temperate setting. The resulting seasonal decomposition identifies the pre-transmission months as the period of highest per-case transmission potential in the Republic of Korea and quantifies the year-crossing transmission structure through which late-season cases seed the following year’s incidence. By relying on routinely collected case, temperature, and vector surveillance data, this approach could be adapted to other temperate *P. vivax* transmission settings with comparable surveillance infrastructure, supporting seasonal targeting of case detection as these countries progress towards elimination.

## Supporting information

**S1 Text. Correction of the pre-2022 malaria vector index for integer-precision rounding**.

**S1 Fig. Distribution of the in-mosquito interval** *I*_*M*_ + *T*_*MH*_. The distribution of *I*_*M*_ + *T*_*MH*_ = *t*_4_ − *t*_2_, the total time the parasite spends within the mosquito, for each combination of *α, β* ∈ {0.2, 0.3, 0.4}.

**S2 Fig. Sensitivity of the annual mean case reproduction number to** *α* **and** *β*. Annual mean of *R*_*t*_ for each year from 2015 to 2023, computed for every combination of *α, β* ∈ {0.2, 0.3, 0.4} (columns, *α*; rows, *β*). The dashed line marks *R*_*t*_ = 1.

**S3 Fig. Sensitivity of the monthly mean case reproduction number to** *α* **and** *β*. Monthly mean of *R*_*t*_ by month, computed for every combination of *α, β* ∈ {0.2, 0.3, 0.4} (columns, *α*; rows, *β*). The dashed line marks *R*_*t*_ = 1.

**S4 Fig. Monthly distribution of cases by intrinsic incubation period**. Mean weekly number of cases per month, separated into cases with a short intrinsic incubation period (*I*_*H*_ < 10 weeks, blue) and a long intrinsic incubation period (*I*_*H*_ ≥ 10 weeks, orange).

## Data and code

All data, analysis code, and materials needed to reproduce the results presented in this study are available at https://github.com/woosikson/malaria-vivax-rt-korea.

## Interactive visualization

Interactive visualization for analyzing the relationship between the symptom onset times of the infector and infectee can be accessed at https://epirt.notion.site/malaria-rt.

## References

1. World Health Organization. World malaria report 2024: addressing inequity in the global malaria response. Geneva: World Health Organization; 2024. Available from: https://www.who.int/publications/i/item/9789240104440.

2. Howes RE, Battle KE, Mendis KN, Smith DL, Cibulskis RE, Baird JK, et al. Global Epidemiology of Plasmodium vivax. American Journal of Tropical Medicine and Hygiene. 2016;95:15–34. doi:10.4269/ajtmh.16-0141.

3. Angrisano F, Robinson L. Plasmodium vivax - How hidden reservoirs hinder global malaria elimination. Parasitology International. 2022;87:102526. doi:10.1016/j.parint.2021.102526.

4. Olliaro PL, Barnwell JW, Barry A, Mendis K, Mueller I, Reeder JC, et al. Implications of Plasmodium vivax Biology for Control, Elimination, and Research. American Journal of Tropical Medicine and Hygiene. 2016;95:4–14. doi:10.4269/ajtmh.16-0160.

5. World Health Organization. Control and elimination of Plasmodium vivax malaria – A technical brief; 2025. Accessed 29 Apr 2025. https://www.who.int/publications/i/item/9789241509244.

6. Byrne I, Cramer E, Nelli L, Rerolle F, Wu L, Patterson C, et al. Characterizing the spatial distribution of multiple malaria diagnostic endpoints in a low-transmission setting in Lao PDR. Frontiers in Medicine. 2022;9:929366. doi:10.3389/fmed.2022.929366.

7. Thriemer K, Ley B, von Seidlein L. Towards the elimination of Plasmodium vivax malaria: Implementing the radical cure. PLoS Medicine. 2021;18:e1003494. doi:10.1371/journal.pmed.1003494.

8. Wallinga J, Teunis P. Different epidemic curves for severe acute respiratory syndrome reveal similar impacts of control measures. American Journal of epidemiology. 2004;160(6):509–16.

9. Huber JH, Johnston GL, Greenhouse B, Smith DL, Perkins TA. Quantitative, model-based estimates of variability in the generation and serial intervals of Plasmodium falciparum malaria. Malaria journal. 2016;15:1–12.

10. Codeço CT, Villela DA, Coelho FC. Estimating the effective reproduction number of dengue considering temperature-dependent generation intervals. Epidemics. 2018;25:101–11.

11. Endo A, Nishiura H. Transmission dynamics of vivax malaria in the Republic of Korea: Effectiveness of anti-malarial mass chemoprophylaxis. Journal of Theoretical Biology. 2015;380:499–505.

12. Kim JH, Lim AY, Cheong HK. Malaria Incidence of the Regions Adjacent to the Demilitarized Zone in the Democratic People’s Republic of Korea, 2004–2016. Journal of Korean Medical Science. 2019;34:e227. doi:10.3346/jkms.2019.34.e227.

13. Lee SY, Lee SD, Oh SK, Park S, Lee JY, Kim J. Introduction to the Second Malaria Re-elimination Action Plan (2024–2028) toward Malaria Elimination by 2030. Public Health Weekly Report. 2024;17(22):962–79. doi:10.56786/PHWR.2024.17.22.3.

14. Korea Disease Control and Prevention Agency. Infectious Disease Portal [Internet] (in Korean); 2026. Weekly notified malaria cases for Seoul, Gyeonggi, Gangwon, and Incheon, obtained from the disease-specific monthly and weekly statistics. Accessed 2026 May 7. Available from: https://dportal.kdca.go.kr/pot/is/rginEDW.do.

15. Korea Meteorological Administration. Open MET Data Portal [Internet] (in Korean); 2026. Daily mean temperatures recorded at the Automated Synoptic Observing System (ASOS) stations in Seoul, Gyeonggi, Gangwon, and Incheon. Accessed 2026 May 7. Available from: https://data.kma.go.kr/climate/RankState/selectRankStatisticsDivisionList.do.

16. Malaria management guidelines 2013 (in Korean). Osong: Korea Disease Control and Prevention Agency; 2013.

17. Malaria management guidelines 2014 (in Korean). Osong: Korea Disease Control and Prevention Agency; 2014.

18. Malaria management guidelines 2015 (in Korean). Osong: Korea Disease Control and Prevention Agency; 2015.

19. Malaria management guidelines 2016 (in Korean). Osong: Korea Disease Control and Prevention Agency; 2016.

20. Malaria management guidelines 2017 (in Korean). Osong: Korea Disease Control and Prevention Agency; 2017.

21. Malaria management guidelines 2018 (in Korean). Osong: Korea Disease Control and Prevention Agency; 2018.

22. Malaria management guidelines 2019 (in Korean). Osong: Korea Disease Control and Prevention Agency; 2019.

23. Malaria management guidelines 2020 (in Korean). Osong: Korea Disease Control and Prevention Agency; 2020.

24. Malaria management guidelines 2021 (in Korean). Osong: Korea Disease Control and Prevention Agency; 2021.

25. Malaria management guidelines 2022 (in Korean). Osong: Korea Disease Control and Prevention Agency; 2022.

26. Malaria management guidelines 2023 (in Korean). Osong: Korea Disease Control and Prevention Agency; 2023.

27. Malaria management guidelines 2024 (in Korean). Osong: Korea Disease Control and Prevention Agency; 2024.

28. Malaria management guidelines 2025 (in Korean). Osong: Korea Disease Control and Prevention Agency; 2025.

29. Gostic KM, McGough L, Baskerville EB, Abbott S, Joshi K, Tedijanto C, et al. Practical considerations for measuring the effective reproductive number, R t. PLoS computational biology. 2020;16(12):e1008409.

30. McKenzie FE, Jeffery GM, Collins WE. Plasmodium vivax blood-stage dynamics. Journal of Parasitology. 2002;88(3):521–35.

31. Ohm JR, Baldini F, Barreaux P, Lefevre T, Lynch PA, Suh E, et al. Rethinking the extrinsic incubation period of malaria parasites. Parasites & vectors. 2018;11(1):1–9.

32. Detinova TS. Age-grouping methods in Diptera of medical importance, with special reference to some vectors of malaria. World Health Organization; 1962.

33. Denlinger DL, Armbruster PA. Mosquito diapause. Annual Review of Entomology. 2014;59:73–93. doi:10.1146/annurev-ento-011613-162023.

34. Mironova V, Shartova N, Beljaev A, Varentsov M, Grishchenko M. Effects of climate change and heterogeneity of local climates on the development of malaria parasite (Plasmodium vivax) in Moscow megacity region. International Journal of Environmental Research and Public Health. 2019;16(5):694. doi:10.3390/ijerph16050694.

35. Feng X, Zhang S, Huang F, Zhang L, Feng J, Xia Z, et al. Biology, bionomics and molecular biology of Anopheles sinensis Wiedemann 1828 (Diptera: Culicidae), main malaria vector in China. Frontiers in Microbiology. 2017;8:1473.

36. Hong H, Eom TH, Trinh TTT, Tuan BD, Park H, Yeo SJ. Identification of breeding habitats and kdr mutations in Anopheles spp. in South Korea. Malaria Journal. 2023;22(1):381.

37. Nah KA, Choi IS, Kim YK. Estimation of the incubation period of P. vivax Malaria in Korea from 2006 to 2008. Journal of the Korean Data and Information Science Society. 2010;21(6):1237–42.

38. Kim SJ, Kim SH, Jo SN, Gwack J, Youn SK, Jang JY. The long and short incubation periods of Plasmodium vivax malaria in Korea: the characteristics and relating factors. Infection & chemotherapy. 2013;45(2):184–93.

39. Kim H, dam Lee S, Shin NR, Hwang K. Status of malaria and diagnosis rate in the Republic of Korea, 2018–2022. Public Health Weekly Report. 2023;16(26):852–66. doi:10.56786/PHWR.2023.16.26.3.

40. Ku B, Shin H, Lee H. Status of follow-up diagnosis after treatment of malaria patients in Republic of Korea, 2014-2020. Public Health Wkly Rep. 2021;14(34):2436–43.

