## Supplementary figures and images for "A time-varying risk assessment framework for *P. vivax* malaria transmission in temperate settings: A case study of the Republic of Korea"

### S1 Fig

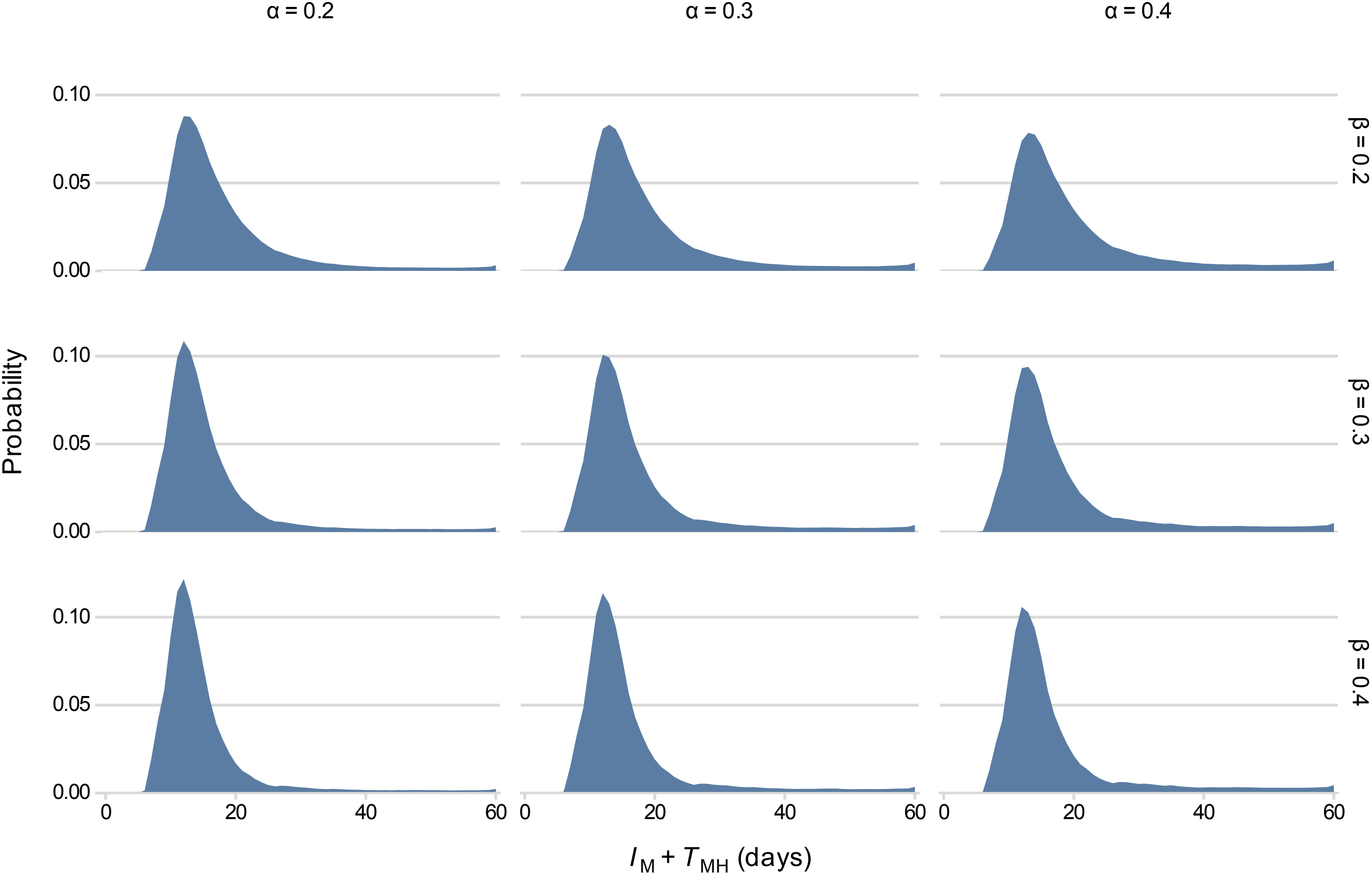

### S2 Fig

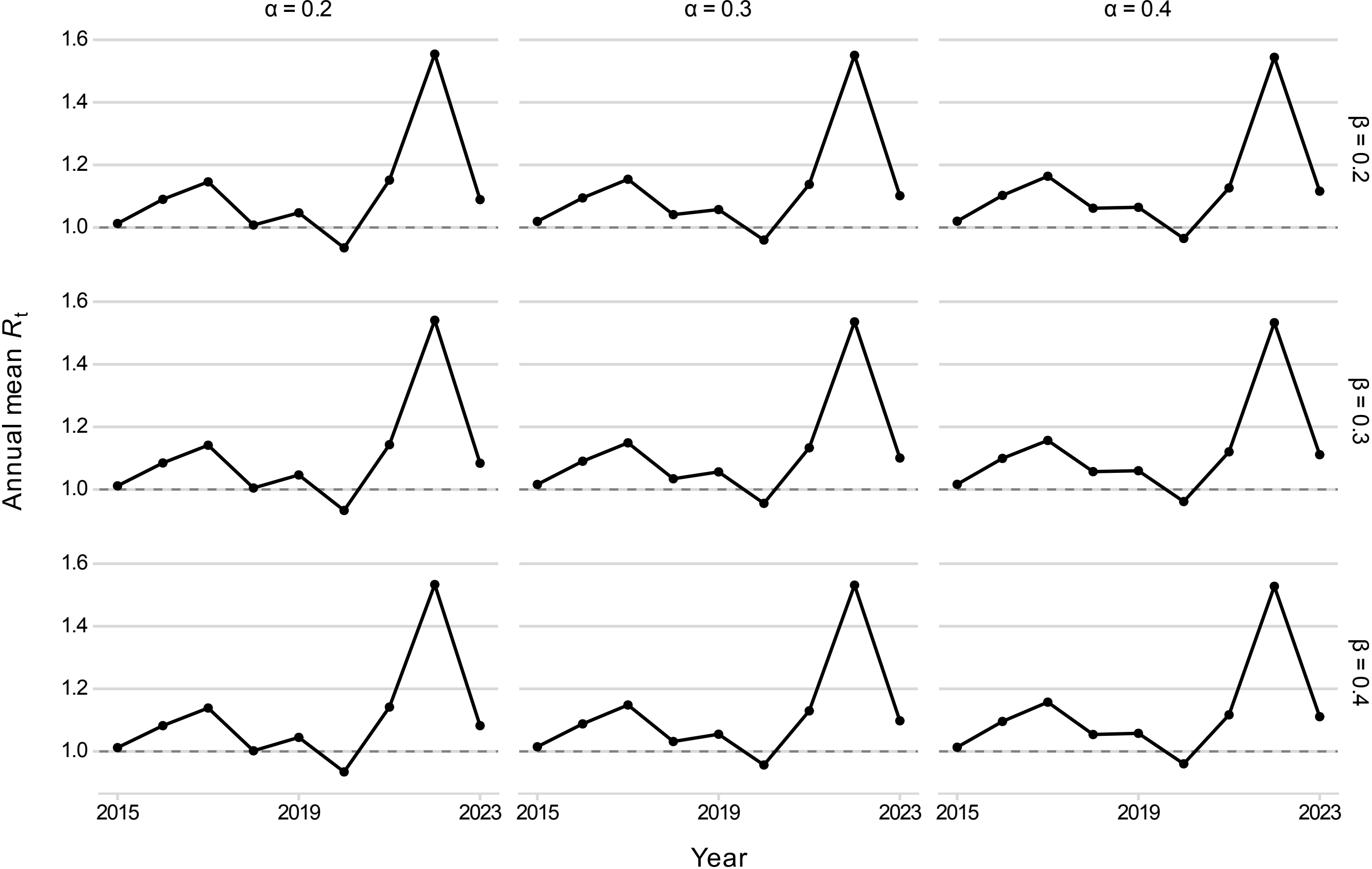

### S3 Fig

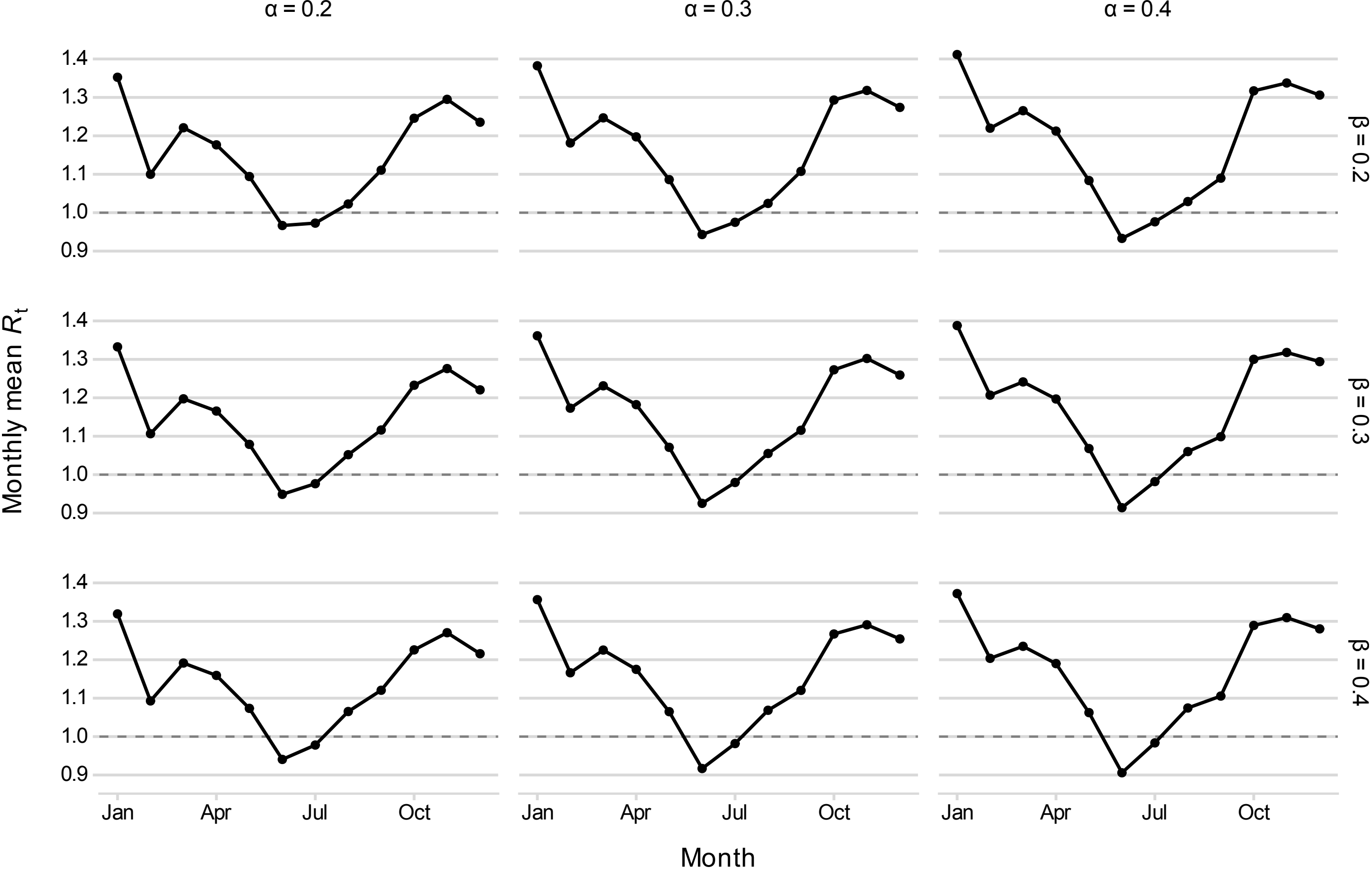

### S4 Fig

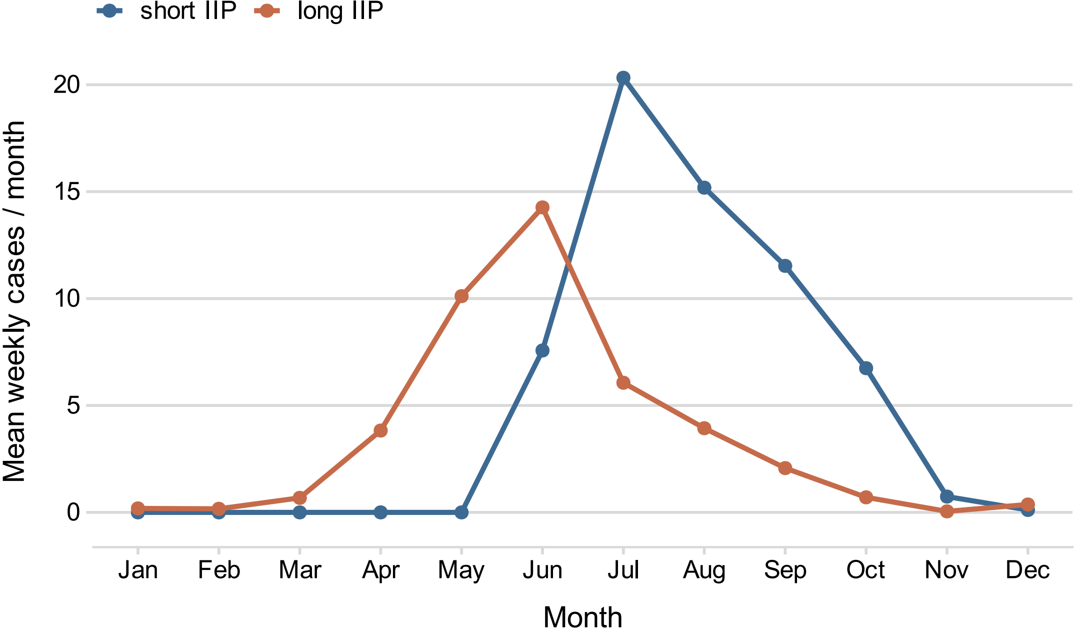
