## Supplementary material for "A time-varying risk assessment framework for *P. vivax* malaria transmission in temperate settings: A case study of the Republic of Korea": S1 Text

**S1 Text. Correction of the pre-2022 malaria vector index for integer-precision rounding.**

The malaria vector index published by the Korea Disease Control and Prevention Agency (KDCA) is, by construction, a non-negative real number that frequently takes small fractional values during low-activity periods. For weeks before 2022, however, the index was recorded only to integer precision: its fractional part was discarded, so that true values below one were entered as 0. Because our transmission model makes human-to-mosquito and mosquito-to-human transmission possible only when the vector index is positive, such a spuriously zero entry would eliminate transmission during weeks when mosquitoes are in fact active, leading to an incorrect estimate of the case reproduction number.

To address this, within the mosquito-active period (weeks 14–44) we replaced vector-index values reported as 0 with 0.1605. This value is the mean of the regional mosquito-surveillance records maintained by the Incheon [1] and Gyeonggi [2] Institutes of Health and Environment over 2013–2022 for precisely those weeks in which the KDCA index was reported as 0; although these records track vector activity only within their respective regions, their finer resolution recovers the sub-unit activity that the pre-2022 national index could not capture.

### References


[1] Incheon Institute of Health and Environment. Malaria vector mosquito surveillance records (in Korean) [Internet]. Incheon: Incheon Institute of Health and Environment [cited 2026 May 7]. Available from: `http://incheon.go.kr/ecopia/index`

[2] Gyeonggi malaria vector mosquito surveillance report 2013–2022 (in Korean). Suwon: Gyeonggi-do Institute of Health and Environment; 2022.
